# Early detection of anastomotic leakage in upper gastrointestinal surgery

**DOI:** 10.1101/2024.05.24.24307864

**Authors:** Felix Merboth, Friederike Sonntag, Katharina Sonntag, Christoph Reißfelder, Daniel E. Stange, Jürgen Weitz, Andreas Bogner

**Affiliations:** Department of Visceral, Thoracic and Vascular Surgery, University Hospital and Faculty of Medicine Carl Gustav Carus, Technische Universität Dresden, Dresden, Germany; National Center for Tumor Diseases (NCT/UCC), Dresden, Germany: German Cancer Research Center (DKFZ), Heidelberg, Germany; University Hospital and Faculty of Medicine Carl Gustav Carus, Technische Universität Dresden, Dresden, Germany; Helmholtz-Zentrum Dresden-Rossendorf (HZDR), Dresden, Germany; Department of General, Visceral and Transplantation Surgery, Medical Faculty Heidelberg, University of Heidelberg, Heidelberg, Germany; Department of Surgery, Universitätsmedizin Mannheim, Medical Faculty Mannheim, Heidelberg University, Mannheim, Germany; General, Visceral and Transplant Surgery, Department of Surgery, Medical University of Graz, Graz, Austria

## Abstract

**Background:** Although surgical methods for upper gastrointestinal tract cancer continue to advance with the aim of reducing the incidence of anastomotic leakage (AL), it remains a prevalent and serious complication. Therefore, early identification of patients at high risk for AL is necessary for timely therapeutic measures.

**Methods:** We retrospectively identified patients with AL who underwent elective gastric/esophageal resection at our department between 2005 and 2017. Using propensity score matching, a comparison group without AL but with comparable baseline characteristics was developed. Several previously published risk scores (o-POSSUM, E-PASS, Steyerberg, NUn score) were calculated, and their predictive accuracy for presence of AL was compared.

**Results:** Steyerberg Risk Score, o-POSSUM, and E-PASS were found to be unsuitable for early detection of an anastomotic problem. However, an increased NUn score on the fourth to seventh postoperative day was independently associated with the presence of AL. The test accuracy (0.631-0.714), sensitivity (28.9%-50.5%), and specificity (72.5%-89.5%) were marginally satisfactory. When only C-reactive protein levels were considered, similar test accuracy (0.629-0.717), sensitivity (47.5%-69.9%), and specificity values (51.5%-81.6%) were observed using a cut-off of 150 mg/l.

**Conclusions:** The NUn score showed no superiority over CRP values in the prediction of AL. Therefore, further diagnostics should be carried out from the fourth postoperative day if the CRP is > 150 mg/l. However, large-scale registry studies and artificial intelligence may aid in more appropriate determination of patient-specific risks in the future.

## Introduction

Cancer of the upper gastrointestinal (GI) tract is among the most common malignancies and the second leading cause of cancer-related death (after lung cancer) globally. Within the framework of multimodal therapy concepts, surgery remains the most relevant component of curative-intent therapies in most cases [1, 2]. However, gastric and esophageal resections are among the most complex oncological surgeries, characterized by high perioperative morbidity and mortality [3]. In recent years, studies have focused on surgical procedures to improve the outcomes. Indeed, minimally invasive and robotic procedures have proven to be beneficial in reducing postoperative complications and shortening of hospital stay [4]. Additionally, the administration of the so-called “selective bowel decontamination” appears to reduce the rate of anastomotic leakage (AL) in upper GI surgery [5].

Despite all improvements, complication rates after gastric or esophageal resection remain high, with AL being the most concerning complication to surgeons. Thus, perioperative scores have been developed to identify patients at risk for postoperative morbidity and mortality. Scores like Steyerberg Risk Score, o-POSSUM, or E-PASS use variables such as age, comorbidity, physiological status, neoadjuvant therapy, hospital volume, surgical factors, and histological features in multilevel logistic regression models to predict postoperative mortality or severity of postoperative complications [6–8]. In contrast to these scores from previous studies, which performed risk stratification based on patient and/or surgical characteristics, other studies have attempted to identify postoperative complications using only laboratory value changes in routine blood samples [9]. Noble et al. developed the NUn score, which is calculated using the log-likelihood ratio of the blood-borne variables albumin, leukocyte count, and C-reactive protein (CRP). Thus, AL can be detected on postoperative day (POD) 4 with high sensitivity and specificity [10].

This retrospective study aimed to compare the predictive power of different perioperative scores for the incidence of AL in gastroesophageal surgery.

## Material and Methods

### Study design

This study was approved by the Institutional Review Board of the Technische Universität Dresden, Dresden, Germany, and was conducted in compliance with the Declaration of Helsinki, according to the ICH Harmonized Tripartite Guideline for Good Clinical Practice (decision number: 224062017). Due to the retrospective study design the ethics committee waived the requirement for informed consent. Before the data were accessed for research purposes at 18.03.2020 all data were fully anonymized. We applied the STROCSS 2021 guidelines, owing to the retrospective nature of our study [11].

We included all patients with esophageal or esophagogastric junction cancer who underwent elective gastroesophageal resection between 2005 and 2017 at the Department of Visceral, Thoracic, and Vascular Surgery of the University Hospital Carl Gustav Carus, Dresden. Esophageal resection types included transhiatal extended gastrectomy, abdomino-thoracic esophagectomy (Ivor Lewis), and abdomino-thoracic-cervical esophagectomy (McKeown). These were performed as open, minimally invasive, robotic, or hybrid procedures. These different surgical procedures were evaluated together because the anastomosis always involves the esophagus and further complications of AL (mediastinitis, empyema, sepsis) and AL treatment strategies (endosponge, drains, esophageal diversion) are the same.

Patient characteristics, preoperative risk factors, and other relevant factors were recorded to calculate the o-POSSUM, E-PASS, and Steyerberg risk scores, according to original publications [6–8]. Serum levels of leukocytes (Gpt/l), CRP (mg/l), and albumin (g/l) within the first 7 postoperative days were used to calculate the NUn score, according to the original formula [10]:

> *NUn score = 11.3894 + (0.005 x CRP) + (0.186 leukocytes) - (0.174 x albumin)*

Postoperative morbidity and mortality rates were evaluated using patient charts and medical records. The severity of postoperative complications was classified using the Clavien–Dindo classification (CDC) [12]. AL definition was in accordance with the 2015 Esophagectomy Complications Consensus Group (ECCG) [13]. In addition, the time of definite AL diagnosis and the diagnostic method used were noted. For this purpose, a contrast leak on computed tomography (CT), endoscopic evidence, or both were scored. Patients were grouped into either AL or non-AL groups, depending on whether they developed AL postoperatively. Subsequently, a 1:2 propensity score matching was performed.

### Statistical analysis and propensity score matching

Owing to the retrospective nature of the present data, the sample size was not selected based on a power calculation. Propensity scores for the AL and control cohorts were calculated using multivariate logistic regression model that included 14 variables (S1 Table). Patients in the two cohorts were matched in a 1:2 ratio, with a maximum difference between propensity scores of 0.05. Continuous variables are expressed as medians and interquartile ranges (IQR) and compared using Student’s t-test or Wilcoxon rank sum test. Dichotomous data were compared using the *χ^2^* test. Variables with p<0.1 in univariate analysis were included in a stepwise backward multivariate logistic regression model. Results were reported as odds ratios (OR) and 95% confidence intervals (CI). The level of significance was set at p<0.05. Receiver operating characteristic (ROC) curves were created to measure the sensitivity and specificity of the scores. The Area Under the Curve (AUC) was measured, and values ranging from 0.6 to 0.7 represented a reasonable test accuracy. Values ˃0.7 represented a good test accuracy. Sensitivity and specificity were determined using ROC curves for the original published NUn score cut-off of 10 and for a separate CRP cut-off. For an optimal CRP cut-off, the value was taken at the point when sensitivity and specificity were close to the AUC value, whereas the absolute difference between sensitivity and specificity was as small as possible [14]. Statistical analyses were performed using IBM SPSS Statistics v23 (IBM Corp., Armonk, NY), and graphical illustrations were performed using GraphPad Prism v7 (GraphPad Software, Inc., La Jolla, CA).

## Results

All patients who underwent esophageal and/or gastric resection for carcinoma in our department from 2015 to 2017 were screened for AL. After performing propensity score matching according to the above mentioned criteria, 146 and 292 patients were classified into the AL and non-AL groups, respectively. Patient characteristics before matching can be found in S2 Table.

### Patient characteristics and surgical findings

Patient characteristics, histopathologic data, and surgical findings did not differ significantly in the two groups on the whole. Only pre-existing pulmonary disease was more frequent in the AL group than in the non-AL group (27.4% vs. 15.1%, p=0.002) and the AL group had markedly more hand-sewn anastomoses (p=0.043). A further detailed list can be found in Table 1.

**Table 1.**
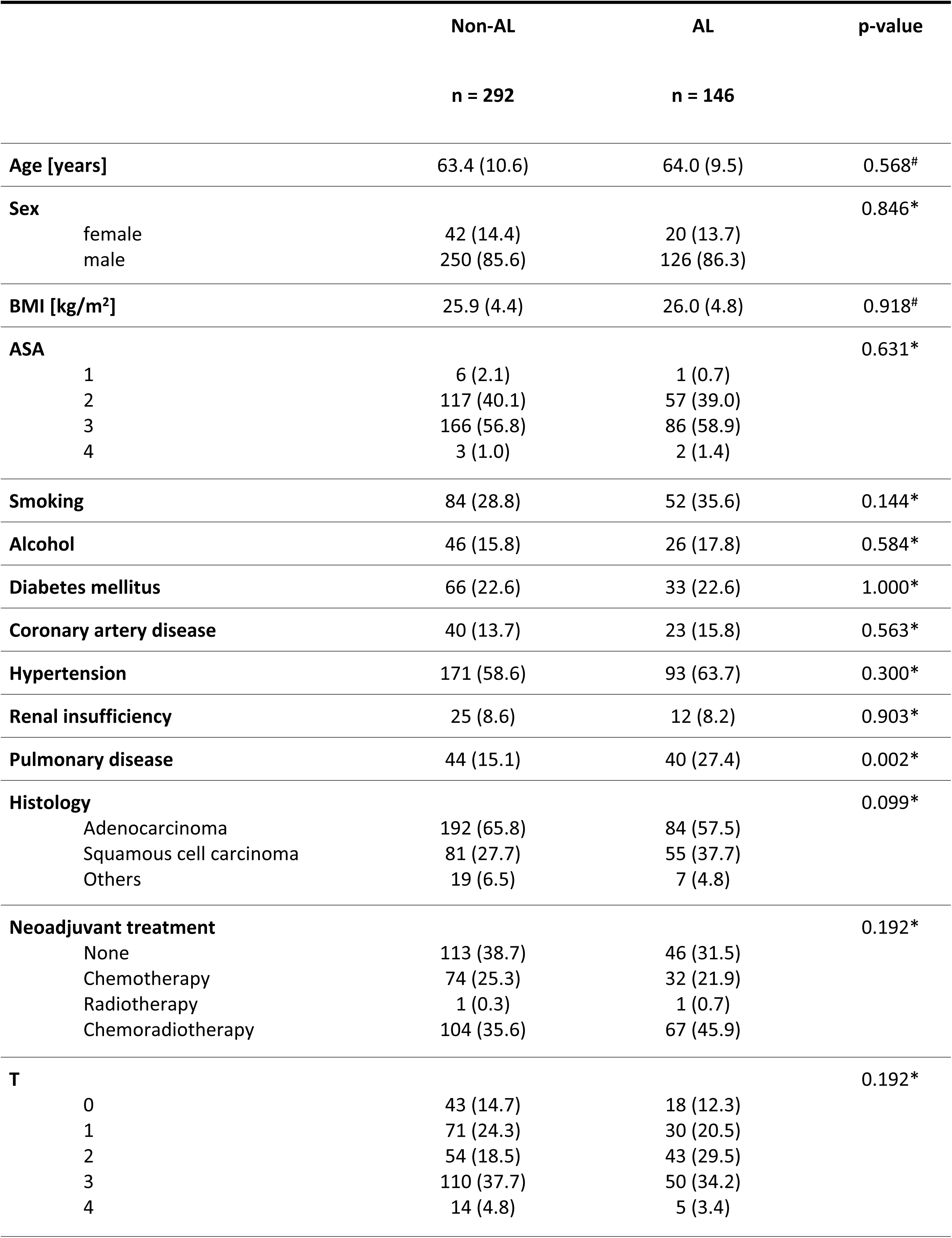
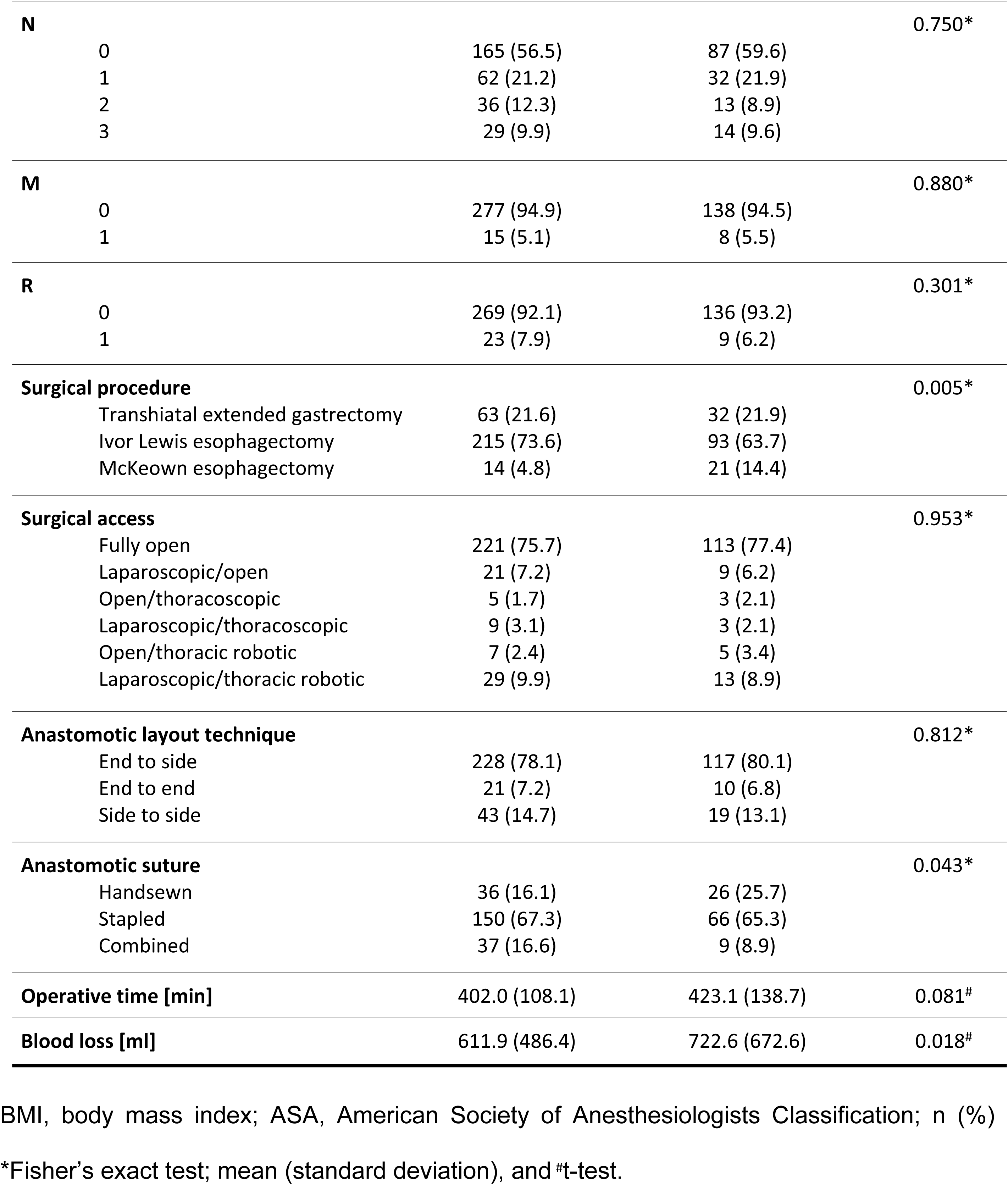
Patient characteristics, histopathologic data, and surgical findings.

All transhiatal extended gastrectomies in both groups (n=95) were reconstructed according to Roux-Y as an end-to-side esophagogastrostomy. For reconstruction after esophageal resection (n=343), a gastric conduit pull-up was used in all patients. In most of these cases, this was performed according to Ivor Lewis with intrathoracic esophagogastrostomy (89.8%). A cervical esophagogastrostomy according to McKeown was used in 10.2%. If the thoracic part was performed minimally invasively, the anastomosis was created side to side with a linear stapler (13.1% vs. 14.7%, p=0.812). In most other cases, the end-to-side anastomosis was created either with a circular stapler (74.7% vs. 77.0%, p=0.448) or by hand suturing (n=5.4% vs. 1.1%, p=0.043). Only in a few cases was an end-to-end anastomosis performed by hand suturing (6.8% vs. 7.2%, p=0.812). The technique of hand-sewn anastomosis depended on the individual surgeon and was therefore not standardized. There were variations between single knot suture, continuous suture or continuous suture of the posterior wall in combination with single knot suture of the anterior wall. All anastomoses were sutured in two rows, and the circular stapler anastomoses were also sutured over.

### Morbidity and mortality

Diagnosis of AL was made according to aforementioned criteria either by contrast leak in CT scan, endoscopically, or both. In the non-AL group, 105 patients (36.0%) had no postoperative complications (CDC=0). Serious complications (CDC≥3a) were observed in 14.0% of patients in the non-AL group and in 69.9% in the AL group (p<0.001). A further detailed list of the CDC distribution is presented in S3 Table. Both 30-d (4.1% vs. 3.1%, p=0.577) and in-hospital mortality (6.8% vs. 6.5%, p=0.892) rates did not differ between the two groups.

More patients in the AL group developed pneumonia during hospitalization (21.5%) than those in the non-AL group (15.0%), and the difference was significant (p=0.007). However, these pneumonias occurred significantly later in the AL group than in the non-AL group (6.3 ± 2.4 d vs. 4.8 ± 2.3 d, p<0.001). In addition, anastomotic insufficiency was diagnosed later than pneumonia, at 10.6 days on average (±13.1 days).

### AL prediction by perioperative scores and laboratory parameters

In univariate and multivariate analyses, o-POSSUM (p=0.320), E-PASS (p>0.692), and Steyerberg Risk Score (p=0.537) did not emerge as risk factors for AL. In contrast, the NUn score was found to be suitable for identifying AL from POD4 (OR 1.526, 95% CI 1.113-2.091, p=0.009), with an increased probability of AL associated with higher NUn scores by POD7 (OR 3.200, 95% CI 1.989-5.150, p<0.001, Table 2). ROC curves were used to determine the diagnostic accuracy of the NUn score (Figure 1). The diagnostic accuracy, expressed by the AUC, was sufficient on the fourth (AUC 0.638) and fifth (AUC 0.631) POD and good on the sixth (AUC 0.714) and seventh (AUC 0.712) POD. Using the NUn score cutoff value of 10 derived from the original publication, the sensitivity and specificity were 36.6% and 80.5% on POD4, which remained unchanged until POD7 (Table 3).

**Figure 1.**
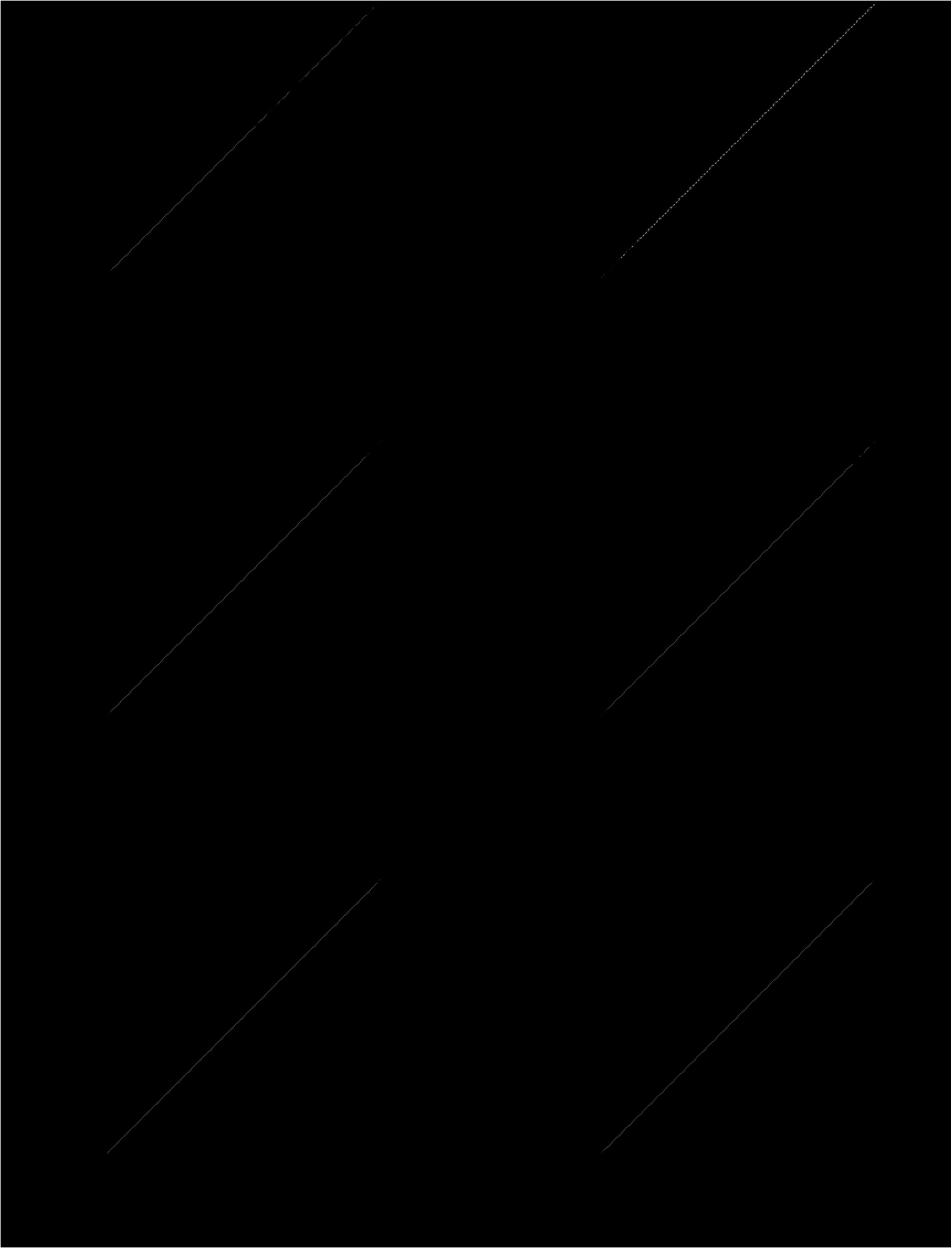
Receiver operating characteristics (ROC) curves and associated area under the curve (AUC) values of the NUn score from postoperative day (POD) 2 to 7

**Table 2.** Univariate and multivariate analysis of Steyerberg, o-POSSUM, E-PASS, and Nun scores.

|  | Univariate |  |  | Multivariate |  |  |
| --- | --- | --- | --- | --- | --- | --- |
|  | Non-AL | AL | p-value | OR | 95% CI | p-value |
| **Steyerberg** | -0.2 (1.6) | 0.2 (1.6) | **0.008** | - | - | 0.537 |
| **o-POSSUM** | 0.3 (0.2) | 0.3 (0.2) | 0.302 | - | - | - |
| **E-PASS** |  |  |  |  |  |  |
| PRS Score | 0.6 (0.3) | 0.7 (0.3) | **0.083** | - | - | 0.692 |
| SSS Score | 0.5 (0.3) | 0.6 (0.3) | **0.067** | - | - | 0.850 |
| CRS Score | 0.8 (0.4) | 0.9 (0.4) | **0.021** | - | - | 0.841 |
| **NUn Score preoperative** | 5.2 (1.1) | 5.3 (1.0) | 0.367 | - | - | - |
| **NUn Score POD1** | 8.4 (1.4) | 8.4 (1.2) | 0.924 | - | - | - |
| **NUn Score POD2** | 8.9 (1.1) | 9.2 (1.2) | **0.029** | - | - | 0.528 |
| **NUn Score POD3** | 9.6 (1.1) | 10.0 (1.1) | **0.008** | - | - | 0.598 |
| **NUn Score POD4** | 9.3 (1.3) | 9.7 (1.2) | **0.002** | **1.526** | **1.113-2.091** | **0.009** |
| **NUn Score POD5** | 9.3 (2.8) | 10.0 (1.1) | **0.010** | **1.906** | **1.322-2.749** | **0.001** |
| **NUn Score POD6** | 8.7 (2.9) | 9.5 (1.2) | **0.003** | **2.732** | **1.790-4.169** | **<0.001** |
| **NUn Score POD7** | 8.8 (1.3) | 9.6 (1.4) | **<0.001** | **3.200** | **1.989-5.150** | **<0.001** |

POD, postoperative day; mean (standard deviation) and t-test; binary logistic regression, OR: odds ratio; 95% CI: 95% confidence interval

**Table 3.**
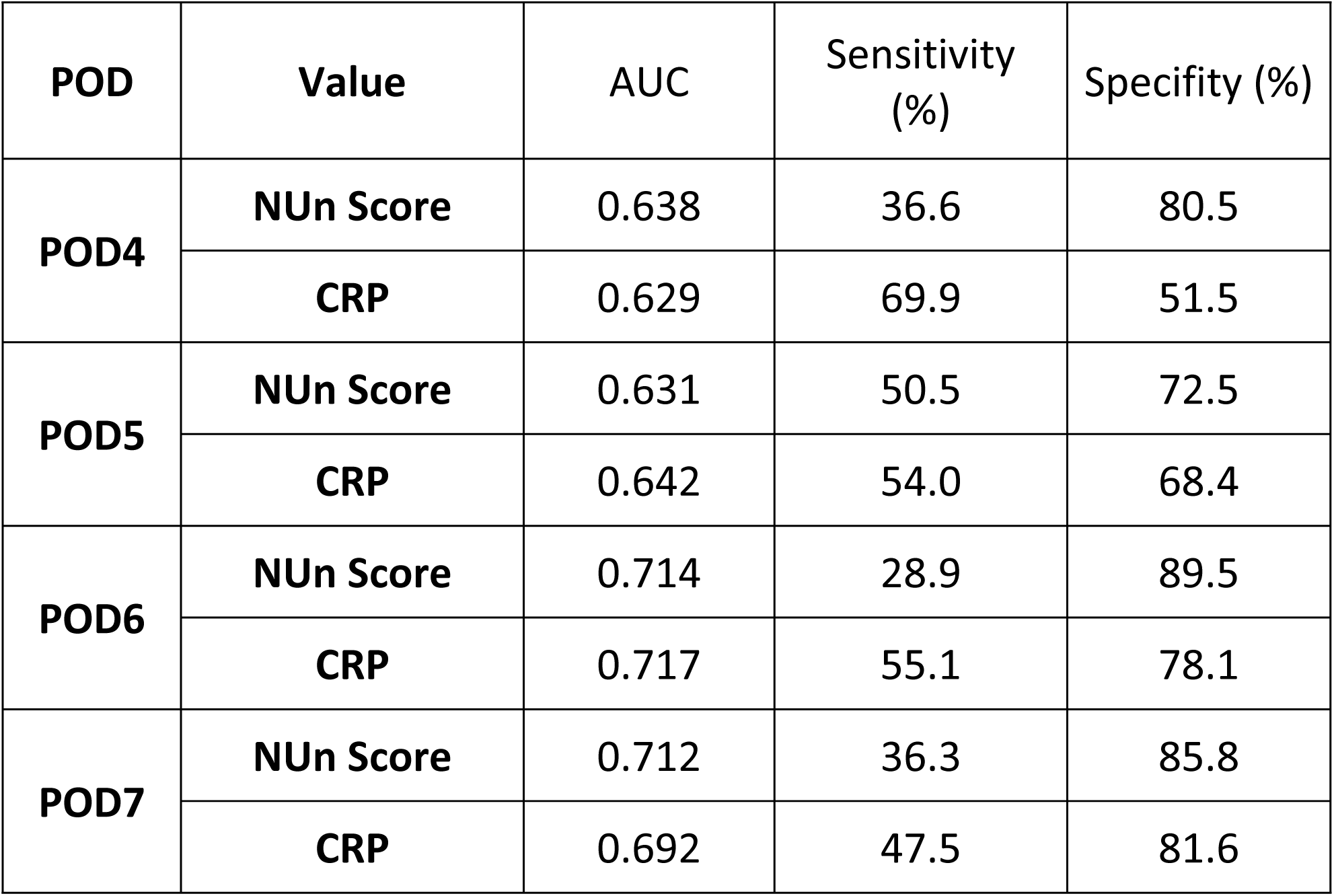
AUC, sensitivity, and specificity within postoperative days 4-7 for NUn score with an original cut-off of 10 and for CRP values with a cut-off of 150 mg/l.

| POD | Value | AUC | Sensitivity (%) | Specificity (%) |
| --- | --- | --- | --- | --- |
| POD4 | NUn Score | 0.638 | 36.6 | 80.5 |
|  | CRP | 0.629 | 69.9 | 51.5 |
| POD5 | NUn Score | 0.631 | 50.5 | 72.5 |
|  | CRP | 0.642 | 54.0 | 68.4 |
| POD6 | NUn Score | 0.714 | 28.9 | 89.5 |
|  | CRP | 0.717 | 55.1 | 78.1 |
| POD7 | NUn Score | 0.712 | 36.3 | 85.8 |
|  | CRP | 0.692 | 47.5 | 81.6 |

However, when the laboratory parameters that constitute the NUn score (leukocytes, CRP, and albumin) were considered individually (Figure 2), large differences in CRP values between the two groups were noted from POD3 (206.3 mg/l vs. 180.7 mg/l, p<0.001) and increased further until POD7 (160.6 mg/l vs. 104.5 mg/l, p<0.001). Leukocytes showed significant differences only from POD5. Albumin levels, on the other hand, were significantly lower in the AL group on POD3 and 4 as well as on POD6 and 7 (S4 Table). Preoperative albumin substitution was generally not carried out. Only postoperative albumin levels below 20.0 g/l were substituted in an adjusted manner.

**Figure 2.**
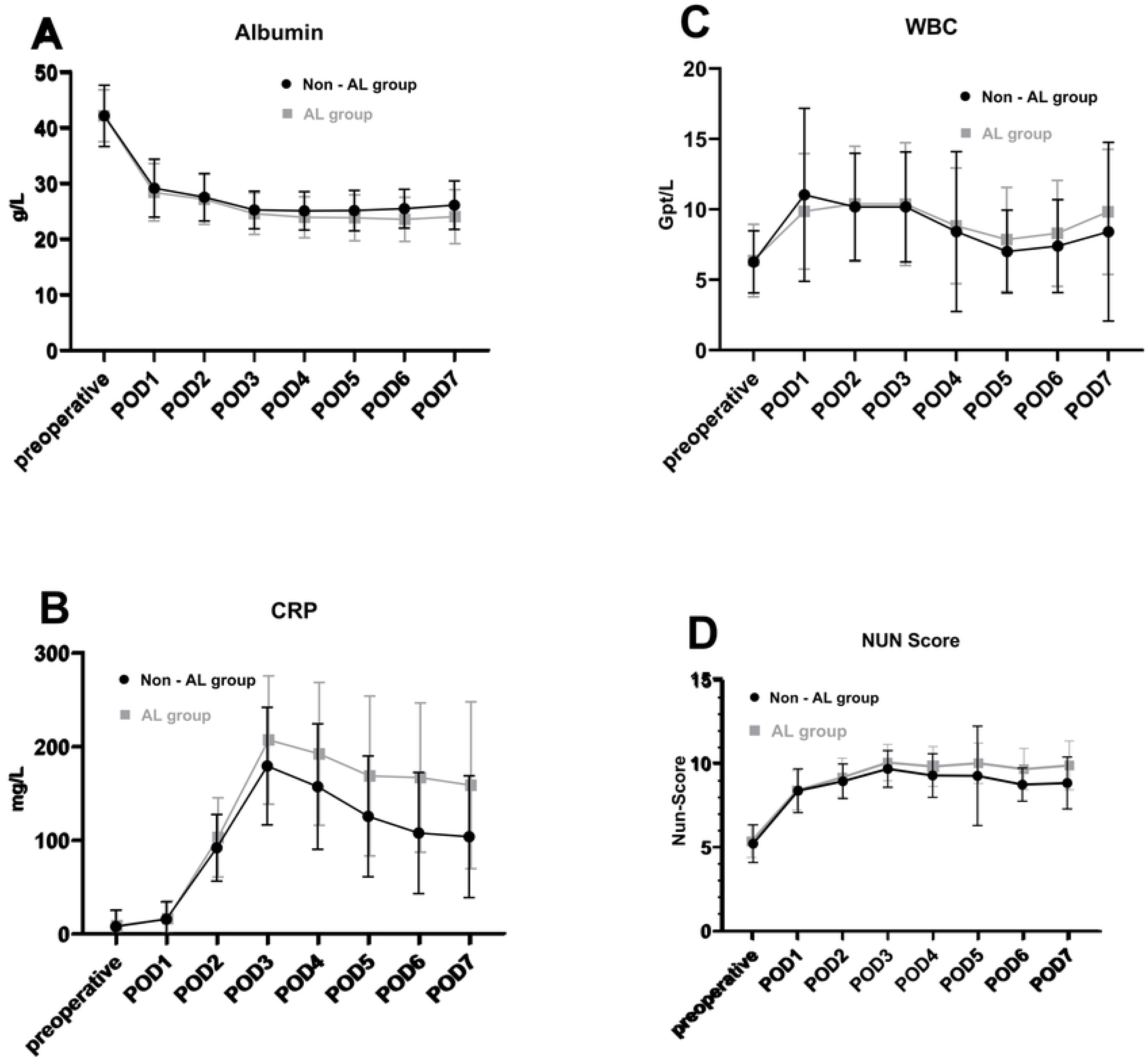
Postoperative course of laboratory parameters a) albumin, b) C-reactive protein (CRP), c) leukocytes (WBC) and the d) NUn score calculated therefrom

ROC curves were created and analyzed for CRP levels (S1 Figure). A CRP level of 150 mg/l was a valid cut-off value. POD4 showed a significantly higher sensitivity (69.9%) with a lower specificity of 51.5%. In the following PODs, however, the sensitivity was marginally higher, with almost equivalent specificity for CRP values compared to the NUn Score (Table 3).

## Discussion

In this retrospective data analysis, perioperative scores were compared in terms of their predictive value for anastomotic insufficiency after esophageal and gastric surgery. To minimize the influence of known patient-related risk factors [9, 15] and instead focus on perioperative factors, a 1:2 propensity score matching for AL with the above-mentioned factors was performed.

Many technical factors influencing the AL rate have been established, including the volume of surgeries conducted by both the hospital and the surgeon, as well as surgical and anastomotic techniques, and the anastomotic site [16–18]. However, patient-specific factors, such as age, obesity, and nicotine consumption have also been identified as risk factors [19]. Surgical and perioperative strategies for gastroesophageal resection have improved in recent years, but nevertheless, an AL rate ˂8% across the board is hardly achievable [20]. In our study, AL was detected in the mean after approximately 11 days. This raises the question of what diagnostic tools can be used to make an early diagnosis and initiate appropriate therapy.

Most risk scores published to date are aimed at short-term outcomes after esophageal and gastric resection rather than AL. Steyerberg Score, o-POSSUM, and E-PASS were slightly increased in the AL group but were not suitable for risk stratification with respect to AL in the multivariate analysis.

In contrast to the previously described scores, the NUn score uses the change in postoperative laboratory values to detect AL. The log-likelihood ratios of CRP, leukocytes, and albumin were used for the calculation. Noble and Underwood, the first two authors after whom the NUn score was named, determined its optimal cut-off as 10 after analyzing the assessment dataset. With this cut-off, a sensitivity of 100%, specificity of 57%, and diagnostic accuracy of 0.879 could be achieved in a validation dataset consisting of 42 patients, including four patients with AL [10]. Other research groups have attempted to apply the NUn score to their cohorts. Bundred et al. showed that in 382 patients (48 of whom had AL), the NUn score significantly predicted AL from POD4. However, the sensitivity, specificity, and diagnostic accuracy achieved were significantly lower at 73%, 65%, and 0.77, respectively [21]. Paireder et al. found lower sensitivity and specificity in their own dataset, resulting in a positive predictive value of only 19.4%. Therefore, the authors concluded that the NUn score was not useful because of its poor discrimination [22]. Findlay et al. found an even poorer test accuracy, with the limitation that their biochemical laboratory had a maximum CRP value of 156 mg/l. Owing to the resulting bias, the group developed their own NUn score cut-off of 7.65 [23]. In contrast to the results of these studies, our dataset had a higher number of patients with AL, with an overall larger sample size. Using the original cut-off of 10, we found a low sensitivity of 37% and acceptable specificity and diagnostic accuracy of 80% and 0.638, respectively. However, the test accuracy of the NUn score in our cohort was far from the value originally described.

A reason for this imprecision could be that the values used for the NUn score are infection-associated parameters that show changes in infectious complications. For example, these parameters may also change in the presence of pneumonia, which is much more common than AL after gastroesophageal resection [3]. The original NUn Score study did not address this complication. However, we were able to show that 21.5% of AL patients and 15.0% of patients in the non-AL group additionally developed pneumonia. In the non-AL group, pneumonia occurred on an average of 5 days postoperatively and was thus exactly within the time frame of use of the NUn score.

Because the formula for calculating the NUn score is rather complicated, other research groups have tried artificial intelligence and machine learning approaches. These new methods can predict overall survival as well as long-term oncological outcomes [24, 25]; nonetheless, the prediction of specific postoperative complications such as that of AL has not been satisfactory. To date, only one research group has shown that CRP-to-albumin ratio and regression tree analysis can achieve a good prediction of AL after esophagectomy [26]. Our objective was to simplify the procedure and expedite decision-making without using complicated formulas or artificial intelligence, in order to achieve greater practicability in everyday clinical practice. Therefore, we examined the test accuracy of only the CRP values. At the determined cut-off of 150 mg/l, the specificity was almost the same as the NUn Score, with marginally better sensitivity (Table 3). The CRP values function more as a search test than as an absolute diagnostic; therefore, the higher sensitivity appears to be advantageous. In the case of increased CRP values from POD4, a secondary examination modality should always be conducted to confirm or rule out AL. Thus, a CT scan of the thorax with oral contrast offers the advantage of detecting both AL and pneumonia [27]. According to the literature, AL is detected late after 9 days on average [28] - in our cohort even after 11 days. Therefore, if CRP is elevated at POD4 and CT findings are ambiguous, endoscopy should be performed and, if necessary, repeated in the following days to exclude or confirm AL with certainty.

## Conclusions

In summary, an increased NUn score from POD4 is suitable for identifying AL. However, the accuracy of the test is unsatisfactory, and the calculation formula is too complicated to be beneficial in routine clinical practice. The test accuracy of postoperative CRP values was nearly equivalent to the NUn Score but these were considerably more manageable. Therefore, we recommend that further diagnostics be performed immediately if the CRP value exceeds 150 mg/l from POD4 for early detection and prompt treatment of AL; this step could potentially prevent devastating complications. However, the cut-off of 150 mg/l determined in our analysis should be transferred to other institutions cautiously since the measurement methods and standard values differ from laboratory to laboratory.

To reliably screen for AL in the postoperative course, new approaches are necessary; artificial intelligence can be used to link laboratory data, radiological diagnostics, and clinical presentations. However, large-scale registry studies are required to obtain sufficient data. Until then, regarding screening for AL, CRP values and clinical appearance should be considered from POD4 to promptly initiate extended diagnostics, such as CT scan or endoscopy, and subsequent treatment.

## Data Availability

All relevant data are within the manuscript and its Supporting Information files.

## Acknowledgements

The authors thank all study nurses, especially Heike Polster, who were involved in this study.

## Abbreviations

AL: Anastomotic leakage
ASA: American Society of Anesthesiologists
AUC: Area Under the Curve
BMI: Body mass index
CDC: Clavien-Dindo Classification
CI: Confidence interval
CRP: C-reactive Protein
CRS: Comprehensive Risk Score
CT: Computed tomography
ECCG: Esophagectomy Complications Consensus Group
E-PASS: Estimation of Physiologic Ability and Surgical Stress
GI: Gastrointestinal
ICU: Intensive care unit
IQR: Interquartile range
OR: Odds ratio
POD: Postoperative day
POSSUM: Physiological and Operative Severity Score for the enUmeration of Mortality and morbidity
PRS: Preoperative Risk Score
ROC: Receiver Operating Characteristics
SSS: Surgical Stress Score

## Supporting information

**S1 Figure.** Receiver operating characteristics (ROC) curves and associated area under the curve (AUC) values of CRP from postoperative day (POD) 2 to 7

**S1 Table.** List of all 14 variables for propensity scoring

**S2 Table.** Patient characteristics, histopathologic data, and surgical findings all patients

**S3 Table.** Morbidity and mortality data of study participants

**S4 Table.** Course of leukocytes, albumin and CRP within the first seven postoperative days.

